# Planned egg freezing over 15 years: return to treatment and success rates in Australia and New Zealand

**DOI:** 10.64898/2026.04.07.26350362

**Authors:** Oisin Fitzgerald, Elena Keller, Peter Illingworth, Devora Lieberman, Michelle Peate, Damian Kotevski, Repon Paul, Iolanda Rodino, Anna Parle, Karin Hammarberg, Tessa Copp, Georgina M Chambers

## Abstract

**Study question:** What are the characteristics and treatment outcomes of women who undertook planned egg freezing (PEF) in Australia and New Zealand between 2009 and 2023?

**Summary answer:** There has been an average yearly increase in the uptake of PEF of 35%, with most women undergoing a single PEF procedure in their mid-thirties. Given ten years follow-up a little over one in four women return, with nearly half of those using donor sperm and one-third achieving a live birth.

**What is known already:** PEF, where women freeze their eggs as a strategy to preserve fertility, has increased dramatically in high income countries in the last decade. Despite the rapid uptake of PEF, there remains limited information to guide women, clinicians and policy makers regarding the characteristics of women undertaking this procedure and treatment outcomes.

**Study design, size, duration:** A retrospective population-based cohort study of all women who undertook PEF in Australia and New Zealand between 2009 and 2023, including their subsequent return to thaw their eggs and treatment outcomes. Where women returned to utilise their eggs, all subsequent embryo transfer procedures were linked enabling calculation of live birth rates per woman.

**Participants/materials, setting, methods:** 20,209 women who undertook PEF in Australia and New Zealand between 2009 and 2023 including 1,657 women who returned to thaw their eggs.

**Main results and the role of chance:** There has been a huge increase in uptake of PEF, from 55 women in 2009 to 4,919 in 2023. Women who freeze their eggs are typically aged 34-38 years (interquartile range) and nulliparous (98.6%). For women with at least 10 years follow-up (i.e. undertook PEF in 2009-13; N=514), 27.9% returned and thawed their frozen eggs (average time to return: 4.9 years). This reduced to 22.1% in those with at least 5 years follow-up (i.e. undertook PEF in 2009-2018; N=4,288). Of those who used their frozen eggs, 47% used donor sperm. After at least two years follow up, 33.9% had a live birth, rising over time to 37.8% for eggs thawed between 2019-2021.

**Limitations, reasons for caution:** In the timeframe 2009-2019 we did not have information on whether egg freezing occurred because of a cancer diagnosis, a cohort we wished to exclude from the study. As a result, for this timeframe we weighted observations by the probability that egg freezing occurred due to cancer, with the prediction model developed on the years 2020-2023.

**Wider implications of the findings:** This study provides recent and comprehensive data on PEF to guide prospective patients and clinicians and inform policy. The exponential growth in PEF in Australia and New Zealand mirrors trends in other high-income countries, suggesting a doubling time of 2-3 years. Study findings highlight the need for setting realistic expectations about the likelihood of returning to use frozen eggs and live birth rates.

**Study funding/competing interest(s):** 2020-2025 MRFF Emerging Priorities and Consumer Driven Research initiative: EPCD000014

## Introduction

Planned egg freezing (PEF), also referred to as ‘elective’ or ‘social’ egg freezing, is a form of assisted reproductive technology (ART), in which a woman undergoes ovarian stimulation to produce multiple mature eggs (oocytes) which are retrieved and then frozen (cryopreserved) for potential future use [1]. In PEF there is no immediate threat to fertility, which is in contrast to medically indicated egg freezing where women with medical conditions such as those requiring gonadotoxic treatment (e.g. chemotherapy) freeze their eggs. As age is the most important factor in determining a woman’s fertility, the intention of PEF is to retrieve eggs at a younger reproductive age and store them for several years, preserving the biological age of the egg [2]. Despite PEF being expensive (around $10-15,000 USD per retrieval cycle) and generally not covered by national health insurance schemes, demand has risen substantially in high-income countries in recent years.

Registry reporting highlights the massive growth in PEF. In the US, information from the Society for Assisted Reproductive Technology (SART) National Summary Report indicates the number of PEF cycles tripled from 13,041 to 39,269 between 2018 and2023 and now represents 10% of non-donor initiated cycles. The Human Fertilisation and Embryology Authority (HFEA) 2023 report, shows the number of PEF cycles in the UK increased from 2,567 in 2019 to 6,932 in 2023, a 170% increase representing 7% of all initiated stimulated treatment cycles. Similarly, the Australia and New Zealand registry reports indicate PEF comprised around 12% of all initiated ovarian stimulation procedures in 2023 [3]. This surge in uptake has generated widespread media coverage and prompted research and debate around issues such as the utilisation rate of frozen eggs [4], the high costs [5], the role of marketing [6], the potentially coercive impact of employers paying women to undertake this procedure, and women accessing their retirement funds to pay for PEF [7].

Despite the rapid uptake of PEF, limited information is available to guide women, clinicians and policymakers regarding the characteristics of women undertaking this procedure, return rates and the chances of having a baby using frozen eggs. To date, most detailed studies, summarised in contemporary reviews [8, 9], rely on data from individual clinics with relatively small sample sizes (<1,000 women) [10-21], raising questions about their generalisability to the wider population. Further, many large national studies (e.g. USA [20, 22-25] and the Netherlands [26]) are missing key information such as the number of eggs frozen or return rates. More importantly, the follow-up time after the initial PEF procedure varies considerably both between studies, often leading to a downward bias in reported return and success rates.

This research aims to answer four inter-related questions based on population-based data from Australia and New Zealand: 1) what are the temporal trends in PEF; 2) what are the characteristics of women who undertake PEF; 3) what are the characteristics of women who return to utilise their frozen eggs, and 4) what are the cumulative? live birth rates?

## Material and methods

### Data and cohort

#### Data

The data for this study comes from the Australia and New Zealand Assisted Reproduction Database (ANZARD), a clinical quality registry comprising information on all ART treatment cycles performed in Australian and New Zealand fertility clinics. This study uses ANZARD data from treatments conducted in 2009-2023. For each individual who underwent PEF, we linked all their subsequent ART cycles to capture the use of any form of ART, either from the previously frozen eggs and/or undertaking a new ART treatment cycle (i.e. ovarian stimulation, egg collection and transfer of any resulting embryos).

The following data items were extracted for all egg freezing cycles at commencement of a PEF cycle: female age at oocyte (egg) pick-up (OPU), female parity at OPU, the number of eggs collected at OPU, the number of eggs frozen. The following data items were extracted at the time that women returned to ART treatment: whether eggs were thawed or a new ART treatment cycle was commenced, source of sperm used for fertilisation via intracytoplasmic sperm injection (ICSI) (ejaculate, epidydimal, testicular), use of donor sperm for ICSI (yes/no), the number of embryos that were transferred and frozen, and clinical pregnancy and live birth outcomes involving the frozen-thawed eggs. Prior to 2020, ANZARD did not record the indication for egg freezing (i.e. planned or for medical indications). For cycles performed between 2020-2023 we additionally extracted the following information: whether the treatment cycle took place for fertility preservation purposes, the reason for female fertility preservation as determined by the treating clinician (cancer diagnosis, other medical reason, or non-medical reason) as well as the sex of the intended parents (female-male, single female, or female-female).

A clinical pregnancy was defined as meeting one of the following: 1) known to be ongoing at 20 weeks; 2) evidence by ultrasound of an intrauterine sac and/or fetal heart; 3) examination of products of conception reveals chorionic villi; or 4) an ectopic pregnancy diagnosed by laparoscopy or by ultrasound. A live birth was defined as the birth of liveborn baby(WHO definition) and of 20 weeks or more gestation or 400 grams or more birthweight [3].

The PEF cohort was defined as: women aged over 18 years with no prior history of ART treatment involving egg collection who underwent an OPU cycle where all eggs were frozen (with no other treatment); the freezing was for fertility preservation purposes and the intention was to store the eggs for more than one year. Women were excluded if the egg freezing was due to cancer treatment or arising as a side-effect of male factor infertility such as a failure to obtain sperm at the time of OPU.

Women meeting these criteria can be explicitly identified using ANZARD data items in cycles occurring in 2020-2023. However, for 2009-2019 cycles the following rules were applied to assign egg freezing cycles to the PEF cohort:

- The eggs were frozen for at least one year
- There was no indication of male infertility (recorded sperm source, male infertility indicator or recorded male age) reported in the egg freezing cycle
- The characteristic of the woman and cycle indicated probabilistically that the treatment did not occur because of cancer treatment. The methodology used to calculate these weights is described below.

Women who return to ART following PEF may either: 1) immediately use their frozen eggs or 2) commence a new ART treatment cycle or 3) a combination of both. When determining the *return rate* we included all these cases and separately note the number/percentage who used their frozen eggs. When calculating descriptive statistics (e.g. age at return to ART) we separated the return cohort into those who returned but never thawed their frozen eggs (e.g. because they achieved a pregnancy using fresh eggs) and those who utilised their frozen eggs. In a small number of cases (N=212) a woman returned to ART and had an embryo freeze-all cycle using fresh eggs. These cases were excluded from our return to ART cohort because there was no attempt at pregnancy.

#### Ethics and reporting guidelines

Ethics approval for this project was obtained from the University of New South Wales Human Research Ethics Committee (correspondence: iRECS0859). We utilised the STROBE reporting guidelines for cohort studies.

#### Data availability statement

The data underlying this study can be made available upon reasonable request to the study authors.

## Analysis

### Weighting by probability of cancer

Using ANZARD data from 2020-2023 we developed a prediction model that predicts whether an egg freezing cycle was undertaken because of cancer treatment. This model was a binomial generalised additive model (logistic link) with binary outcome cancer (yes/no) and predictors (all recorded at the freezing cycle): female age, infertility diagnosis, number of eggs retrieved, number of eggs frozen, number of freezing cycles, and year of treatment initiation. In both the training (2020-2023) and 2009-2019 datasets, the cohorts included were PEF and cancer related egg freezing, with other egg freezing cycles (e.g. male infertility) removed. This model was then used to predict the probability *p* that a woman in the 2009-2019 cohort was undertaking egg freezing due to cancer, with each woman then getting the weight *w* = 1 − *p*. For 2020-2023 women the weights are deterministic, i.e. *w* = 0 if the relevant “reason for egg freezing” data item indicated the treatment was related to cancer treatment and the women is excluded from the PEF cohort (as noted above).

### Descriptive statistics

Categorical variables are presented as numbers (percentage), while continuous variables are presented as either median [25th percentile; 75th percentile] or mean (standard deviation) depending on whether they are skewed. Categorical variables are presented graphically using barplots (percentage). Continuous variables are presented as histogram or where comparison between strata or grouping variable (e.g. year of freeze) is required as density plots (2 groups) or boxplots (>2 groups). Differences between distributions are tested using the Kolmogorov-Smirnov test. Our definition of live birth rate is:

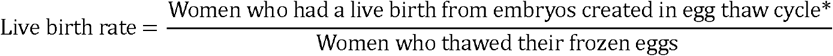

*In some cases (33.7%) there is a concurrent fresh OPU and egg thawing, in which case the data source cannot distinguish whether an embryo was created from fresh or frozen-thawed eggs.

Live birth rates are calculated excluding thaw cycles that occurred in 2022 or 2023 (i.e. 2009-2021) giving women at least one year follow-up (no live birth data is available for 2023). Along with the full 2009-2021 cohort, live birth rates are reported only for thaw cycles that occurred in 2019-2021 to give a sense of the most contemporary outcomes. For age group (18-34 years, 35-39 years and 40-44 years) calculations, we only use 2019-2021.

### Software

The analysis is performed using R version 4.5, utilising the packages *targets, tableone, ggplot2, mgcv* and *data*.*table*.

## Results

### Overview of cohort

As shown in Table 1, between 2009 and2023, 20,209 women (weighted) undertook PEF in Australia and New Zealand. On average (mean), they underwent 1.5 OPU cycles, freezing 12 [IQR: 7; 18] eggs. Most (68%) had one OPU cycle, 22% had two and 10% had three or more OPU cycles. Nearly all women were nulliparous (98.6%, excluding those missing this information).

**Table 1.** Characteristics of women who engaged in planned egg freezing (PEF) in Australia and New Zealand, 2009-2023.

|  | All women | Women who returned to ART treatment but did not thaw their frozen eggs | Women who thawed their frozen eggs |
| --- | --- | --- | --- |
| **Patient and treatment characteristics at initial egg freezing cycle** |  |  |  |
| Number of women (weighted) | 20,209 | 667 | 1,657 |
| Female age (years) at initial OPU | 35.6 [33.6; 37.7] | 36.2 [34.6; 38.1] | 37.3 [35.5; 39.0] |
| Intending parent sex at initial OPU (2020-2023 only) |  |  |  |
| *Female-male* | 5.0% | 5.6% | 4.4% |
| *Female-female* | 0.8% | 0.4% | 1.2% |
| *Single female* | 94.1% | 93.9% | 94.4% |
| Parity at initial OPU |  |  |  |
| *Parous* | 1.3% | 0.9% | 1.5% |
| *Nulliparous* | 93.7% | 89.3% | 88.3% |
| *Missing* | 4.9% | 9.8% | 10.2% |
| Number of egg freezing cycles per woman | 1.5 (0.9) | 1.6 (1.0) | 1.7 (1.1) |
| Egg freezing method |  |  |  |
| *Slow freezing* | 1.2% | 2.2% | 4.1% |
| *Vitrification* | 98.8% | 97.8% | 95.9% |
| Total number of eggs collected per woman | 14 [9; 22] | 13 [8; 20] | 14 [8; 22] |
| Total number of eggs frozen per woman | 12 [7; 18] | 10 [6; 15] | 11 [6; 18] |

At study follow-up, 2,324 women (weighted) had returned to ART treatment following PEF. Of these 1,657 (71.3%) thawed their frozen eggs, with the other 28.7% using fresh eggs created from a new ART cycle. Table 1 outlines the characteristics of the full cohort (weighted) stratified by whether the woman used their frozen eggs or fresh eggs. Comparing those who returned to ART (either to use their frozen eggs or start a new ART cycle) with the full cohort of women who undertook PEF, those who returned tended to be older at egg freezing and had collected and frozen marginally fewer eggs at OPU. Comparison of the distributions of number of eggs collected and frozen between those who returned and those who did not, suggested a statistically significant difference (*p*<0.01).

As shown in Table 2 most women who thawed their frozen eggs were close to 40 years, with 50% of thaws being in women aged 38.7-42.6 years. Approximately 43.5% of egg thaws were undertaken by single women. Relatedly nearly half (47.0%) of fertilisation procedures using thawed eggs used donor sperm. One in five (21.7%) women did not thaw all their eggs, with the number thawed eggs per woman 10 [IQR: 6; 14] compared to the 11 [IQR: 6; 18] frozen. One third (33.9%) of women having their first egg thaw in 2009-2021 had at least one live birth using thawed eggs, with 2.4% having more than one live birth. These rates improved with time, with 37.8% of women undertaking their first egg thaw in 2019-2021, having at least one live birth using thawed eggs, and 4.7% having more than one live birth.

**Table 2.** Overview of women who returned to assisted reproductive technology after planned egg freezing (PEF) in Australia and New Zealand, 2009-2023.

|  | Women who started a new ART cycle (without thawing their frozen eggs) | Women who thawed their frozen eggs (with or without starting a new ART cycle) |
| --- | --- | --- |
| **Patient and treatment characteristics at first treatment cycle upon return to ART** |  |  |
| Number of women (weighted) | 667 (28.7) | 1,657 (71.3%) |
| Female age at treatment reinitiation | 38.5 [36.3; 40.1] | 40.7 [38.7; 42.6] |
| Intending parent sex (2020-2023 only) |  |  |
| *Female-male* | 46.9% | 53.5% |
| *Female-female* | 1.4% | 3.0% |
| *Single female* | 51.7% | 43.5% |
| Use of donor sperm |  |  |
| Yes | 45.3% | 47.0% |
| No | 49.6% | 52.4% |
| *Not applicable\* or missing* | 4.8% | 0.5% |
| Male partner age (female-male couples) | 39.3 [36.1; 42.4] | 40.9 [37.0; 44.7] |
| Total number of eggs thawed per woman | - | 10 [6; 14] |
| Women who started a new ART cycle while thawing eggs | - | 34.5% |
| *Number of eggs collected per woman* | - | 7 [4; 11] |
| *Total number of fresh and frozen-thaw eggs per woman* | - | 15 [10; 24] |
| *Outcomes using frozen-thawed eggs (2009-2021)* |  |  |
| At least one live birth per woman<sup>†</sup> | - | 33.9% |
| More than one live birth per woman<sup>††</sup> | - | 2.4% |
| Live birth rate per embryo transfer | - | 26.0% |
| *Outcomes using frozen-thawed eggs (2019-2021)* |  |  |
| At least one live birth per woman<sup>†</sup> | - | 37.8% |
| More than one live birth per woman<sup>††</sup> | - | 4.7% |
| Live birth rate per embryo transfer | - | 29.5% |

\*Cases where no eggs were suitable for fertilisation
\*\*An embryo that was transferred during a fresh cycle or frozen for later transfer
\*\*\*Transferred all available fresh and frozen embryos - the data source does not track embryo disposal
<sup>†</sup>Calculated using thaw cycles that occurred before 2021 to allow at least two years follow-up.
<sup>††</sup>Calculated using thaw cycles that occurred before 2019 to allow at least four years follow-up.

### Characteristics of egg thaw cycles

Figure 2 shows the return rates and outcomes of the thaw cycle and resulting embryos amongst the 1,657 women (weighted) who returned to thaw their frozen eggs in 2009-2023. Given at least 5 years follow-up (i.e. undertook PEF in 2009-18; N=4,288), 22.1% of women who undertake PEF return to thaw their frozen eggs, typically after 4.4 [IQR: 3.1; 5.5] years (Figure 2A and 2B). Given at least 10 years follow-up (i.e. undertook PEF in 2009-13; N=514), 27.9% of women who undertake PEF return to thaw their frozen eggs, typically after 5.3 [IQR: 3.7; 7.1] years. A larger percentage of women returned to any form of ART (whether a new stimulated cycle and/or egg thawing) – 29.2% after 5 years and 37.9% after 10 years. Figure 2C shows the distribution of age at egg freezing and thaw, with the typical women in this cohort freezing their eggs at 37.3 years and thawing them at 40.7 years. As shown in Figure 2D, very few women have total egg thawing failure with >99% of women who thawed eggs in 2019-2021 having at least one egg fertilise. From there 97% have at least one day 1 embryo, 86% have an embryo suitable for transfer or freezing, 81% undertake an embryo transfer, and 37.8% have a live birth. The live birth rate per women using frozen-thawed eggs is graphed in Figure 2D, showing the expected decline around 35 years at the time of egg freezing.

### Trends in planned egg freezing

Figure 3 illustrates trends in PEF over time. As shown in Figure 3A there has been a 35% per annum increase in the number of women undertaking PEF since 2009. There has been an associated increase in the number of eggs in frozen storage (Figure 3B). Note that the data source does not record egg disposal, meaning the true number in storage as of December 2023 may have been lower (Figure 3B). The age at freezing has gradually decreased since 2009, with more women in their early to mid-thirties freezing their eggs in recent years (Figure 3C). While the number of women thawing their eggs is growing exponentially (35% per year), the absolute numbers are still relatively small, with just over 500 women thawing their eggs in 2023 (Figure 3D). Just under 400 babies have been born from eggs frozen as part of PEF since 2009 (Figure 3E). There has been a trend towards improving live birth rates per woman using frozen eggs over time, with rates of 36.4-39.7 % per year for cycles using eggs from thaws initiated since 2019-2021 (Figure 3F).

## Discussion

This study characterises women who undertook planned egg freezing (PEF) in Australia and New Zealand between 2009-2023, including trends in the procedure across this timeframe and the outcomes for women who returned and thawed their frozen eggs. This is one of the largest and most contemporary studies of PEF to date and data can be used to inform decision making and clinical counselling. Key findings include a yearly growth in PEF of 35% and women typically freezing 7-18 eggs at age 34-38 years (interquartile ranges (IQRs). Given at least 10 years follow-up, 27.9% thaw their frozen eggs, with 47% using donor sperm, and 33.9% achieving a live birth, rising to 37.8% for eggs thawed between 2019-2021.

The number of eggs available for fertilisation is a key predictor of success in all forms of assisted reproductive technology (ART). As noted, we found that most women are freezing around 12 [IQR: 7; 18] eggs, having undertaken one (68%) or maybe two (22%) PEF cycles (Figure 1B). However, four of the five egg freezing calculators reviewed by Wolf, Minis [27] agree that for a 33 year old woman to have a high (74-97%) chance of a live birth she must freeze at least 20 eggs. Presumably factors such as the high financial cost of the procedure, physical burden of treatment, indirect costs from time off work, and the tendency for people to over-estimate ART success rates [28] play a role in many freezing fewer eggs than optimally needed for a high chance of success according to these calculators. The resulting live birth rate per woman of 37.8% (2019-2021) suggests that PEF as currently practiced is an uncertain form of “fertility insurance”, and advertising slogans such as “start a family later without worrying about your biological clock” [6] are misleading.

**Figure 1.**
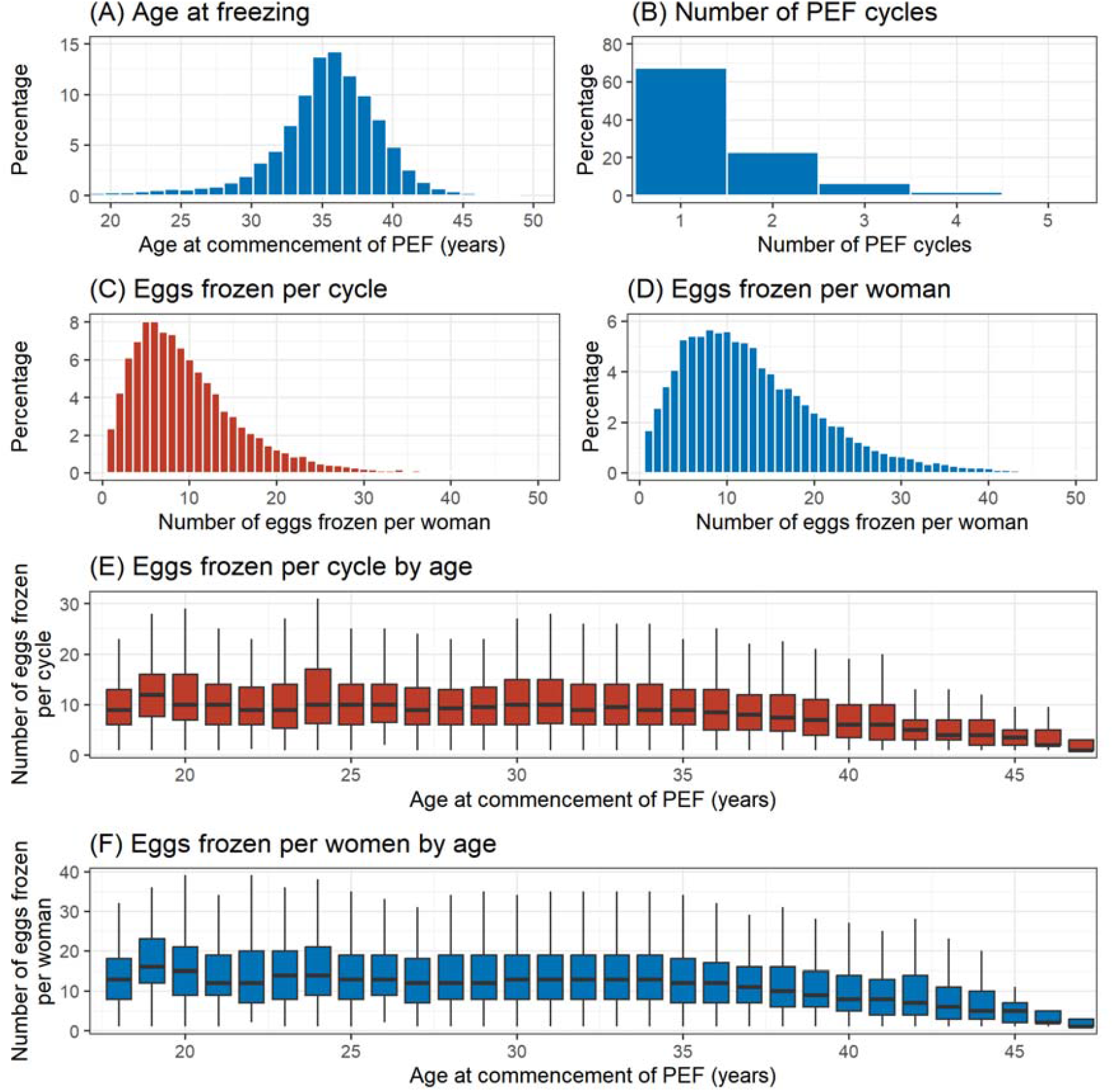
Characteristics of planned egg freezing (PEF) cycles, Australian and New Zealand, 2009-2023. (A) Age of women at commencement of first PEF cycle. (B) Number of PEF cycles undertaken per woman. (C) Number of eggs frozen per PEF cycle. (D) Number of eggs frozen per woman undertaking PEF. (E) Number of eggs frozen per PEF by female age at commencement of first PEF cycle. (F) Number of eggs frozen per woman by female age at commencement of first PEF cycle.

Another key predictor of ART success is the age of the woman at egg retrieval, with egg freezing (vitrification) preserving the reproductive capacity of the egg. On average, ART success is relatively stable until 35 years after which it declines rapidly to around a 1-2% chance of a live birth per egg retrieval procedure by the mid-forties (YourIVFSuccess Estimator). We found that half of women are freezing their eggs in their mid-thirties (34-38 years), with 59% over 35 years. The average age at freeze has been declining over time (Figure 3C), which will contribute towards improved success rates of PEF over time. This trend has been observed in other studies [29], suggesting increasing awareness that freezing at an earlier age is important. However, given the high cost of egg storage, and relatively low return rates (Figure 1A), it is important women considering egg freezing receive accurate information around the relationship between age and decline in egg quality (e.g. Figure 2E). Indeed, existing evidence suggests that women who freeze at earlier ages are less likely to return to use them [25], potentially leading to difficult decisions around what to do with surplus frozen eggs [30].

**Figure 2.**
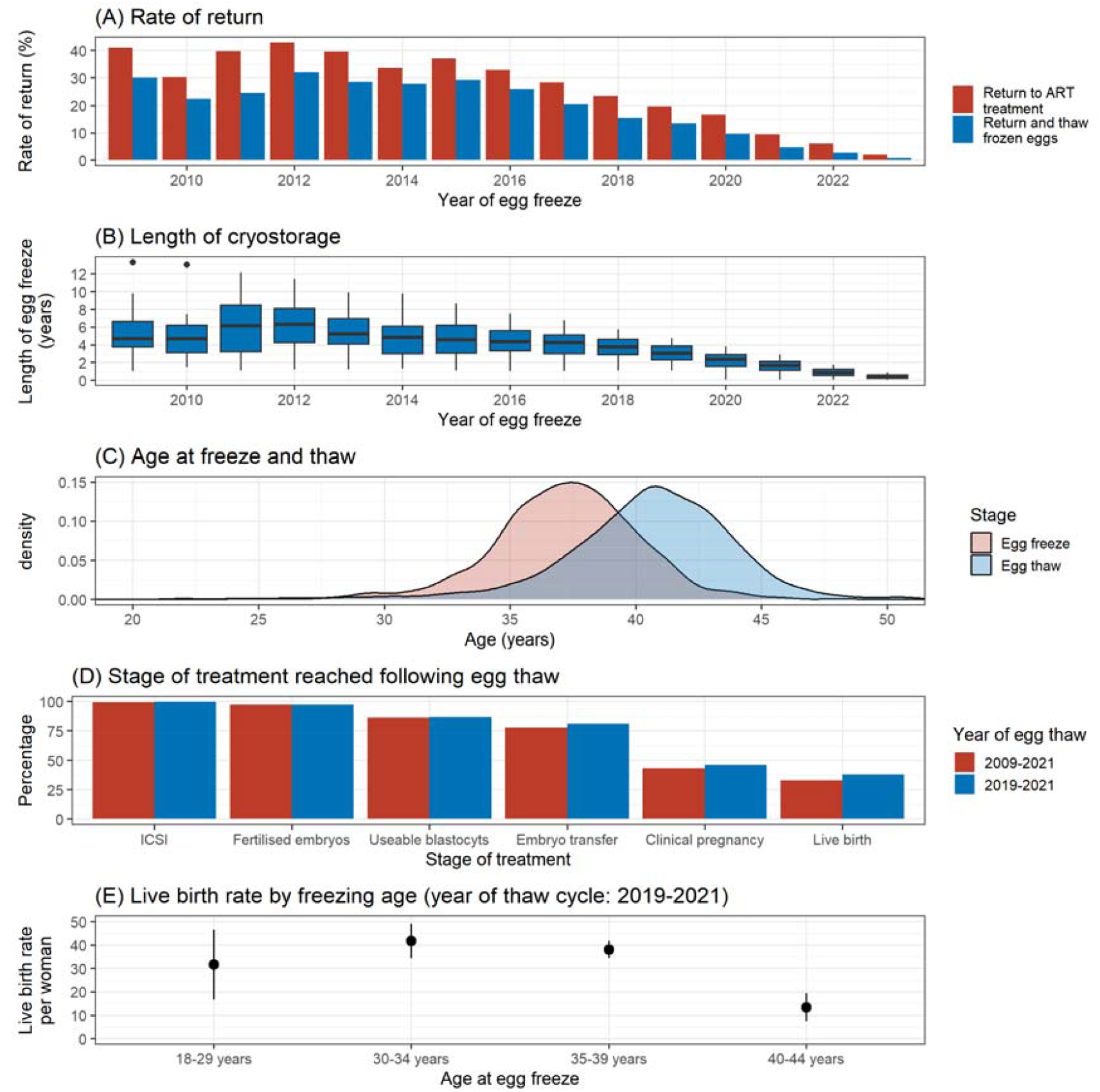
Characteristics of egg thaw cycles following planned egg freezing (PEF), Australia and New Zealand, 2009-2023. (A) Percentage of women who use their frozen eggs by year of first PEF cycle. (B) Length of egg freeze amongst women who use their frozen eggs by year of initiation of first PEF cycle. (C) Age at egg freeze and thaw of women who thawed their frozen eggs. (D) Stage of ART treatment reached by women using frozen-thawed eggs stratified by timeframe (egg thaw in 2009-2021 or 2019-2021) and given at least one year follow-up. (E) Live birth rate per woman using frozen-thawed eggs by female age for those who thawed their eggs in the timeframe 2019-2021 giving at least two years follow-up.

**Figure 3.**
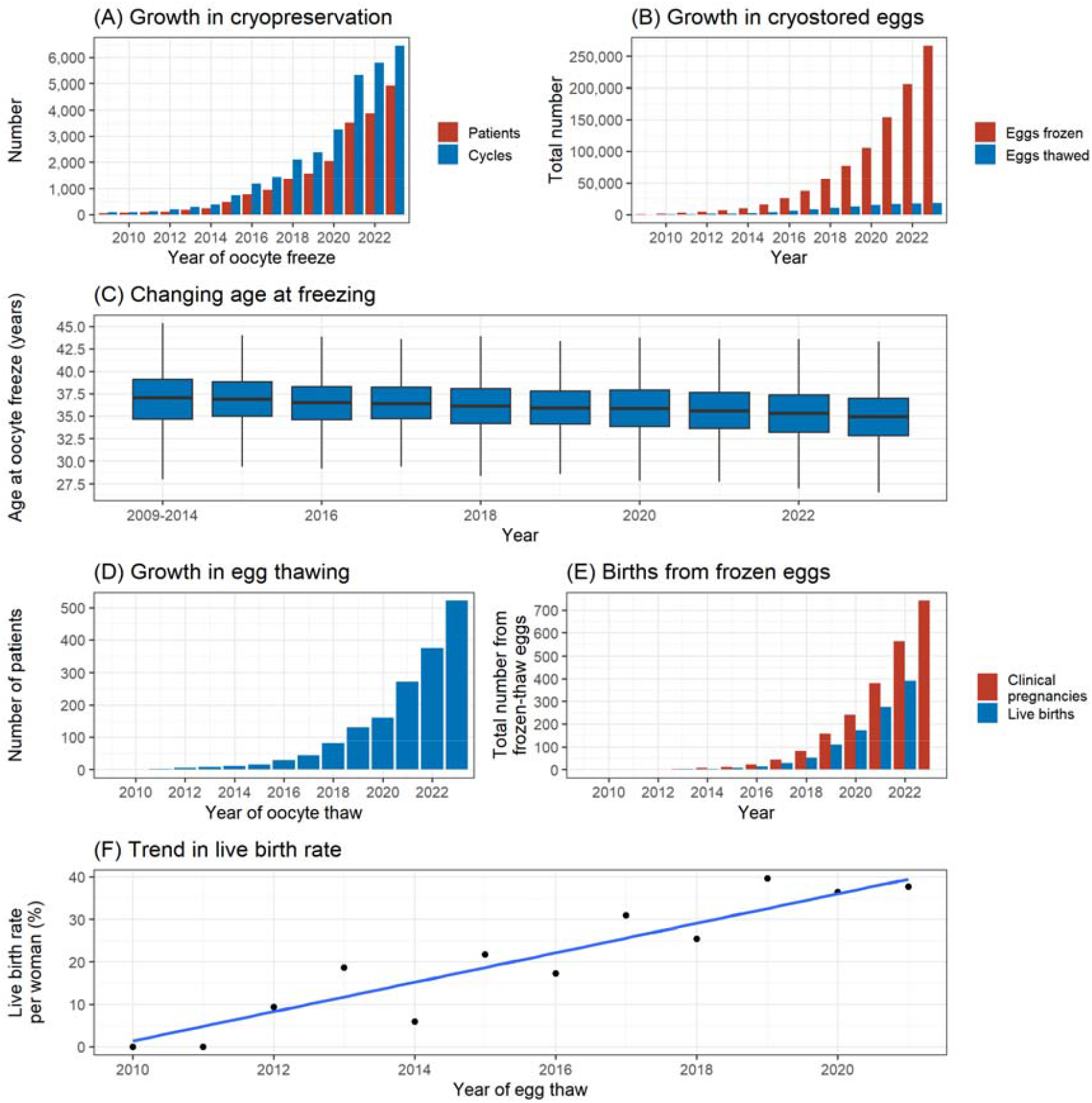
Trends in planned egg freezing (PEF) procedures and associated thaw cycles, Australia and New Zealand, 2009-2023. (A) Number of women undertaking PEF cycles per year. (B) Total (cumulative) number of eggs that have been frozen and thawed since 2009. (C) Trend in distribution of female age at initiation of PEF by year. (D) Number of women thawing their eggs frozen as part of PEF per year. (E) Total (cumulative) number of clinical pregnancies and live births that resulted from eggs frozen as part of a PEF cycle since 2009. (F) Trend in live birth rate per woman from ART cycles using frozen-thawed eggs.

Given 10 years follow-up, just over one quarter (27.9%) of women return to use their frozen eggs following PEF, having stored them for 4.9 [IQR: 3.3; 6.6] years. Previous literature has reported a wide range of return rates for women who undertake PEF, varying from 2.5-38.1% [4, 10, 13, 19, 31, 32]. However, variation in follow-up time adds ambiguity around whether the return rates are truly different across these cohorts, or just a reflection of different follow-up periods. A recent meta-analysis found 10.8% of women returned to use their frozen eggs after a median follow-up time of 7 years [9]. It appears clear that to date, much less than half of women return to use their frozen eggs.

This study is not without limitations. In the timeframe 2009-2019 we did not have information on whether egg freezing occurred because of a cancer diagnosis, a cohort we wished to exclude from the study. As a result, for this timeframe we weighted observations by the probability that egg freezing occurred due to cancer, with the prediction model developed on the years 2020-2023. In the current study we did not investigate the impact of age at egg thaw on reproductive outcomes. In prior adjusted analyses we have found declining live birth rates in women using donated eggs once the egg recipient is aged over 44-45 years [33], with further research needed to investigate whether a similar effect exists for age at egg thaw.

PEF is often promoted as a new reproductive option that empowers women by stopping the biological clock and allowing them to have children beyond their most fertile years [34]. Whilst PEF offers a strategy to potentially circumvent age related infertility, it is paramount that women are informed about the merits and limitations of PEF. This includes discussions about treatment outcomes such as the low return rates, the number of eggs required for a reasonable chance of a live birth and the risks associated with pregnancy at advanced maternal age, and the psychosocial factors that influence the decision. This can be achieved through pre-treatment counselling which incorporates discussions about all aspects of PEF and possible alternative pathways to parenthood and the use of empirically based decision aids that assist women in making informed and value congruent decisions about if and when to freeze their eggs [35].

In conclusion, the exponential growth in PEF has established it as a mainstream ART treatment. It accounted for 12% of initiated ART treatment cycles in Australia and New Zealand in 2023. PEF is often portrayed as an insurance against future infertility, however, as confirmed in this study, it does not provide a guaranteed path to parenthood. The results of this study, provide crucial information to guide women, clinical and counselling practice and inform policy.

## Data Availability

The data underlying this study can be made available upon reasonable request to the study authors.

